## Supplementary material for "Measures to assess quality of postnatal care: a scoping review": S2_Text

**Final Pubmed (457)**

("postnatal care"[MeSH Terms] OR "postnatal care"[Title/Abstract] OR "post partum care"[Title/Abstract] OR "postpartum care"[Title/Abstract] OR "post partum period"[Title/Abstract] OR "postpartum period"[Title/Abstract] OR "postnatal period"[Title/Abstract] OR "puerperium"[Title/Abstract] OR "perinatal period"[Title/Abstract] OR "perinatal care"[Title/Abstract]) AND ("Health Status Indicators"[MeSH Terms] OR "indicators"[Title/Abstract] OR "indicator"[Title/Abstract] OR "index"[Title/Abstract] OR "assessment"[Title/Abstract] OR "assessments"[Title/Abstract] OR "measurement"[Title/Abstract] OR "measurements"[Title/Abstract] OR "measure"[Title/Abstract] OR "measures"[Title/Abstract]) AND ("quality of care"[Title/Abstract] OR "coverage"[Title/Abstract] OR "experience of care"[Title/Abstract] OR "provision of care"[Title/Abstract] OR "adequate care"[Title/Abstract] OR "evidence based care"[Title/Abstract] OR "patient-centered care"[Title/Abstract] OR "efficient care"[Title/Abstract] OR "respectful care"[Title/Abstract] OR "timely care"[Title/Abstract] OR "effective care"[Title/Abstract] OR "equitable care"[Title/Abstract] OR ("safe care"[Title/Abstract] OR "quality care"[Title/Abstract] OR "content of care"[Title/Abstract] OR "package of care"[Title/Abstract] OR "high quality"[Title/Abstract] OR "self-care"[Title/Abstract] OR "selfcare"[Title/Abstract] OR "self-care"[Title/Abstract])) AND (2010/01/01:2022/12/31[Date - Publication] NOT ("animals"[MeSH Terms] NOT "humans"[MeSH Terms]))

**Final Embase (496)**

(('perinatal care'/exp OR 'postnatal care'/exp OR 'postpartum'/exp OR 'puerperium'/exp) AND 'postnatal care':ti,ab,kw OR 'post partum care':ti,ab,kw OR 'postpartum care':ti,ab,kw OR 'post partum period':ti,ab,kw OR 'postpartum period':ti,ab,kw OR 'postnatal period':ti,ab,kw OR 'puerperium':ti,ab,kw OR 'perinatal period':ti,ab,kw OR 'perinatal care':ti,ab,kw) AND ('measurement'/exp OR 'indicator'/exp OR indicators:ti,ab,kw OR indicator:ti,ab,kw OR index:ti,ab,kw OR assessment:ti,ab,kw OR assessments:ti,ab,kw OR measurement:ti,ab,kw OR measurements:ti,ab,kw OR measure:ti,ab,kw OR measures:ti,ab,kw) AND ('quality of care'/exp OR 'quality of care' OR coverage:ti,ab,kw OR 'experience of care':ti,ab,kw OR 'provision of care':ti,ab,kw OR 'adequate care':ti,ab,kw OR 'evidence based care':ti,ab,kw OR 'patient-centered care':ti,ab,kw OR 'efficient care':ti,ab,kw OR 'respectful care':ti,ab,kw OR 'timely care':ti,ab,kw OR 'effective care':ti,ab,kw OR 'equitable care':ti,ab,kw OR 'safe care':ti,ab,kw OR 'quality care':ti,ab,kw OR 'content of care':ti,ab,kw OR 'package of care':ti,ab,kw OR 'high quality':ti,ab,kw OR 'self care':ti,ab,kw OR 'selfcare':ti,ab,kw OR 'self-care':ti,ab,kw) AND [2012-2022]/py

### Final Scopus (934)

(TITLE-ABS-KEY("postnatal care" OR "post partum care" OR "postpartum care" OR "post partum period" OR "postpartum period" OR "postnatal period" OR "puerperium" OR "perinatal period" OR "perinatal care") AND TITLE-ABS-KEY("Health Status Indicators" OR "indicators" OR "indicator" OR "index" OR "assessment" OR "assessments" OR "measurement" OR "measurements" OR "measure" OR "measures") AND TITLE-ABS-KEY("quality of care" OR "coverage" OR "experience of care" OR "provision of care" OR "adequate care" OR "evidence based care" OR "patient-centered care" OR "efficient care" OR "respectful care" OR "timely care" OR "effective care" OR "equitable care" OR "safe care" OR "quality care" OR "content of care" OR "package of care" OR "high quality" OR “self care” OR “selfcare” OR “self-care” )) AND ( LIMIT-TO ( PUBSTAGE,"final" ) ) AND ( LIMIT-TO ( PUBYEAR,2022) OR LIMIT-TO ( PUBYEAR,2021) OR LIMIT-TO ( PUBYEAR,2020) OR LIMIT-TO ( PUBYEAR,2019) OR LIMIT-TO ( PUBYEAR,2018) OR LIMIT-TO ( PUBYEAR,2017) OR LIMIT-TO ( PUBYEAR,2016) OR LIMIT-TO ( PUBYEAR,2015) OR LIMIT-TO ( PUBYEAR,2014) OR LIMIT-TO ( PUBYEAR,2013) OR LIMIT-TO ( PUBYEAR,2012) OR LIMIT-TO ( PUBYEAR,2011) OR LIMIT-TO ( PUBYEAR,2010) ) AND ( LIMIT-TO ( DOCTYPE,"ar" ) OR LIMIT-TO ( DOCTYPE,"re" ) ) AND ( EXCLUDE ( SUBJAREA,"BIOC" ) OR EXCLUDE ( SUBJAREA,"AGRI" ) OR EXCLUDE ( SUBJAREA,"IMMU" ) OR EXCLUDE ( SUBJAREA,"PHAR" ) OR EXCLUDE ( SUBJAREA,"NEUR" ) OR EXCLUDE ( SUBJAREA,"ARTS" ) OR EXCLUDE ( SUBJAREA,"BUSI" ) OR EXCLUDE ( SUBJAREA,"VETE" ) OR EXCLUDE ( SUBJAREA,"COMP" ) OR EXCLUDE ( SUBJAREA,"DECI" ) OR EXCLUDE ( SUBJAREA,"EART" ) OR EXCLUDE ( SUBJAREA,"ECON" ) OR EXCLUDE ( SUBJAREA,"MATH" ) OR EXCLUDE ( SUBJAREA,"DENT" ) )

### Final Cinahl (441)

(MH "Perinatal Nursing") OR (MH "Perinatal Care")) OR (MH "Postnatal Period+") OR (MH "Postnatal Care+") OR (MH "Puerperium") OR  MH "Perinatal Nursing") OR (MH "Perinatal Care") OR "postnatal care" OR "post partum care" OR "postpartum care" OR “post partum period”  OR “postpartum period”  OR “postnatal period”  OR "puerperium" OR “perinatal period”  OR “perinatal care”)

AND

"indicators" OR "indicator" OR "index" OR "assessment" OR "assessments" OR "measurement" OR "measurements" OR "measure" OR “measures”

AND

“quality of care”  OR “coverage” OR “experience of care” OR “provision of care” OR “adequate care” OR “evidence based care” OR “patient-centered care” OR “efficient care” OR “respectful care” OR “timely care” OR “effective care” OR “equitable care” OR “safe care” OR “quality care” OR “content of care” OR “package of care”  OR “high quality” OR “self care”

+ Manual filters regarding data and publication type

**Final Web of Science (415)**

((TS=("postnatal care" OR "post partum care" OR "postpartum care" OR “post partum period” OR “postpartum period” OR “postnatal period” OR "puerperium” OR “perinatal period” OR “perinatal care”)) AND TS=("Health Status Indicators" OR "indicators" OR "indicator" OR "index" OR "assessment" OR "assessments" OR "measurement" OR "measurements" OR "measure" OR “measures”) AND ALL=(“quality of care” OR “coverage” OR “experience of care” OR “provision of care” OR “adequate care” OR “evidence based care” OR “patient-centered care” OR “efficient care” OR “respectful care” OR “timely care” OR “effective care” OR “equitable care” OR “safe care” OR “quality care” OR “content of care” OR “package of care” OR “high quality” OR “selfcare” OR “self-care” OR “self care”))

+Add manual time restriction
